# Application of the Weight Self-Stigma Questionnaire (WSSQ) in Subjects with Overweight and Obesity living in Italy

**DOI:** 10.1101/2022.02.22.22271322

**Authors:** Quintiliani Livia, Sisto Antonella, Vicinanza Flavia, Bertoncini Ilaria, Valentina Pasquarelli, Manfrini Silvia, Watanabe Mikiko, Tuccinardi Dario, Curcio Giuseppe

## Abstract

**Aims:** Although weight-based stigmatization is widespread in everyday life, a suitable measure of weight self-stigma is currently unavailable for those with overweight/obesity whose primary language is Italian. The purpose of this study was to translate and test the psychometric properties of the Italian version of the Weight Self-stigma Questionnaire (I-WSSQ) to administer it to adults with overweight/obesity.

**Materials and Methods:** A cross-sectional study was conducted on 214 adults with overweight or obesity. Data including Body Mass Index (BMI), I-WSSQ, and Body Uneasiness Test (BUT) scores were collected from April the 2^nd^ to July the 6^th^, 2021.

**Results:** Reliability was assessed through internal consistency: the Cronbach α of the I-WSSQ was α =.814, indicating good reliability. The exploratory factor analysis revealed a two-factor structure of I-WSSQ (self-devaluation and fear of enacted stigma, respectively) that explained 49,25% of the total variance. In addition, the I-WSSQ score directly correlated with BMI and BUT score, indicating an acceptable criterion-related validity.

**Conclusion:** The I-WSSQ shows adequate reliability and validity. Health professionals may use the I-WSSQ to assess the weight self-stigma of adults with overweight/obesity whose primary language is Italian.

## Introduction

Obesity is a significant public health problem worldwide, with numbers rapidly growing in Italy as well ^1,2^. Not only weight excess leads to physical complications, some of which are well established ^3-6^ and some others are emerging^7,8^, but it also substantially affects the psychological^9,10^, emotional ^11^ and social dimensions ^12,13^. The simple imbalance between energy intake and expenditure explains a small part of the obesity pandemic we face, and several other factors, such as endocrine disruptors, epigenetics, infections, increased maternal age, and medications, may play a significant role ^14-16^. However, to date, the pathogenesis of obesity is far from being fully elucidated, leading to the frequent misconception that weight gain is solely due to the “bad” habits of the individual. In fact, the lives of those with obesity may be disrupted by the multiple forms of prejudice and discrimination to which they are exposed ^17-19^: obesity is often seen as dependent on internal factors which are directly controllable, such as the absence of physical activity, overeating, and, more generally, the lack of willpower ^20,21^, with the consequence that these people are often pointed out as lazy, lacking in intelligence and self-control, less competent and not compliant with medical care and with poor self-discipline ^22-24^. These stereotypes generate the stigma of obesity and influence social interactions, resulting in a significant psychosocial and depression-related stressor in high-income countries ^24,25^. Therefore, in recent years, the concept of social stigmatization and its influence on medical and psychological research has gained increasing interest ^26-28^.

Sikorski and colleagues ^29^ have highlighted how the attribution of obesity to internal factors is the primary source of stigmatization and discrimination of subjects suffering from this condition, leading to less empathy and willingness to support them. The internalization of stigmatizing attitudes and behaviours even by the individual concerned or family members increases the emotional distress, leading to social isolation and psychological deterioration ^30,31^. Candidates to bariatric surgery also suffer from the same stigma: in a recent study, 87% of bariatric surgery candidates reported being stigmatized due to the choice of undergoing surgery, a decision seen as lack of willpower and “cheating”, as opposed to losing weight with more effort through diet and exercise. As a result, over half of the interviewed subjects reported hiding the surgery from at least one person ^32,33^. Unfortunately, these pervasive stereotypes contribute to prejudice and discrimination in many areas of daily life, such as employment, healthcare, education, and the media.

Thus, the availability of recent, high-quality data on health-related stigma or self-stigma is critical to improve interventions and programs addressing them, yet such data are often lacking, also due to a lack of assessment tools validated in languages other than English ^34^. Moreover, patients with obesity living in Italy may differ from those living in English speaking countries for the cultural environment, dietary habits (i.e. Mediterranean diet) and psychosocial characteristics, and self-stigma therefore needs to be evaluated in this specific population with appropriate tools. The Weight Self-Stigma Questionnaire (WSSQ) estimates weight-related self-assessment and stigmatization of people with obesity^35^. However, it is currently not validated in Italian, making it not applicable to this population.

We therefore aimed to translate the WSSQ into Italian (I-WSSQ) and test its psychometric properties to validate its use in individuals with overweight/obesity whose primary language is Italian.

## Methods

### Methodology

The WSSQ was translated from English into Italian through the following steps: *Preparation*: we contacted the questionnaire’s creator, obtaining written permission to use and translate the tool. *Forward translation*: two freelance translators individually translated the WSSQ into Italian (I-WSSQ). *Reconciliation*: the I-WSSQ was reviewed, and disagreement was reconciled through an expert panel discussion. *Back translation*: the I-WSSQ was back-translated into English by a third translator. Then, two expert panel members reviewed the back-translated I-WSSQ to guarantee the conceptual equivalence of the wording ^36^.

The local ethics committee approved this study (Prot.: PAR 31.21 (OSS) 31/03/2021). Written informed consent was obtained from all participants prior to enrolment. All the procedures performed in the study complied with the Helsinki Declaration.

### Participants

Two hundred fourteen patients whose primary language is Italian, with a Body Mass Index (BMI) ≥25 Kg/m^2^, were enrolled in the study and recruited through consecutive random enrolment lasted three months (from April to July 2021).

Structured questionnaires were used to collect sociodemographic and anthropometric data (age, gender, and the I-WSSQ and Body Uneasiness Test (BUT) were administered in person. The BMI was measured as weight in kilograms divided by the square of height in meters by registered dietitians.

### Instruments/ Measures

The current study involved the administration of the self-report questionnaires listed below:

#### *Italian version of the Weight Self-stigma Questionnaire* (I-WSSQ)

As the WSSQ, the I-WSSQ comprises a 12 item Likert-type scale that measures weight-related self-stigma. It consists of two subscales of 6 elements each for measuring weight-related self-devaluation (item number 1 to 6) and fear of enacted stigma (item number 7 to 12), with no reverse-scored items ^35^. The I-WSSQ asks participants to rate their perception of stigma from others, ranging from 1 (completely disagree) to 5 (completely agree). Total scores are calculated for the two subscales ranging from 12 to 60, with higher scores indicating higher weight-related self-devaluation and fear of enacted stigma.

#### Body Uneasiness Test (BUT)

The BUT is a multidimensional self-administered questionnaire for the clinical assessment of body uneasiness in subjects suffering from eating disorders and/or obesity^37^. It consists of two parts: the first part (BUT-A) includes 34 items, and scores are combined into a global severity index (GSI) and five subscales that evaluate weight phobia, concern for body image, compulsive control, avoidance behaviours and the sense of detachment and depersonalization towards one’s body (e.i. Item 1 - *I spend a lot of time in front of the mirro*r, or Item 10 - *I make detailed comparisons between my appearance and that of others*); the second part (BUT-B) includes 37 elements and scores are combined into two global measures (total positive symptoms (PST) and positive symptom distress index (PSDI) and eight factors that evaluates specific concerns regarding various parts of the body and other related aspects such as concerns related to one’s smell, redness, sweat and noises produced by one’s body (e.i. *of my body, in particular, I hate. Item 3-The shape of my face)*. The score is distributed on a 6-point Likert scale from 0 to 5, where 0 corresponds to never and 5 to always; the sum corresponds to the total BUT score. Higher scores indicate greater physical discomfort.

In the original validations study, the levels of Cronbach’s alpha coefficients range between .64 and .89.

### Data Analysis

Similar to the original WSSQ validation process ^2^, the internal consistency of the I-WSSQ was assessed using the Cronbach α coefficient. Analyses of the correlation between the scale items were conducted using the Kaiser-Meyer-Olkin sample adequacy measure and the Bartlett test to assess the suitability of exploratory factor analysis. An exploratory factor analysis using principal factor extraction was performed to examine the factorial structure of the scales, the oblique rotation method was used to obtain clear factorial structures and enable comparison with the original study results ^33^; moreover, factor extraction was done with eigenvalues above 1, while estimation method applied was the minimum residual. Finally, also a confirmatory factor analysis has been run.

Finally, a criterion-related validity was examined by performing a correlation analysis between the BMI, BUT-A (GSI TOT and DISTRESS), BUT-B (PST and DISTRESS), and the I-WSSQ. There were no missing data. All statistical tests were performed using the SPSS statistical package (version 25). The statistical significance for the analyses was set at p ≤ .05.

## Results

### Characteristics of study participants

A total of 214 participants participated in this study: the descriptive statistics of the sample are displayed in **Table 1**. Briefly, 61 males (28.5%) and 153 females (71.%) were enrolled, with an age between 18 and 74 years old, average 46 ± 11 years old. Among these, 15 (7%) had a BMI between 25 and 29, 19 (8.9%) between 30 and 35, 73 (34.1%) between 36 and 40 and 107 (50%) with a BMI > 40.

**Table 1.**
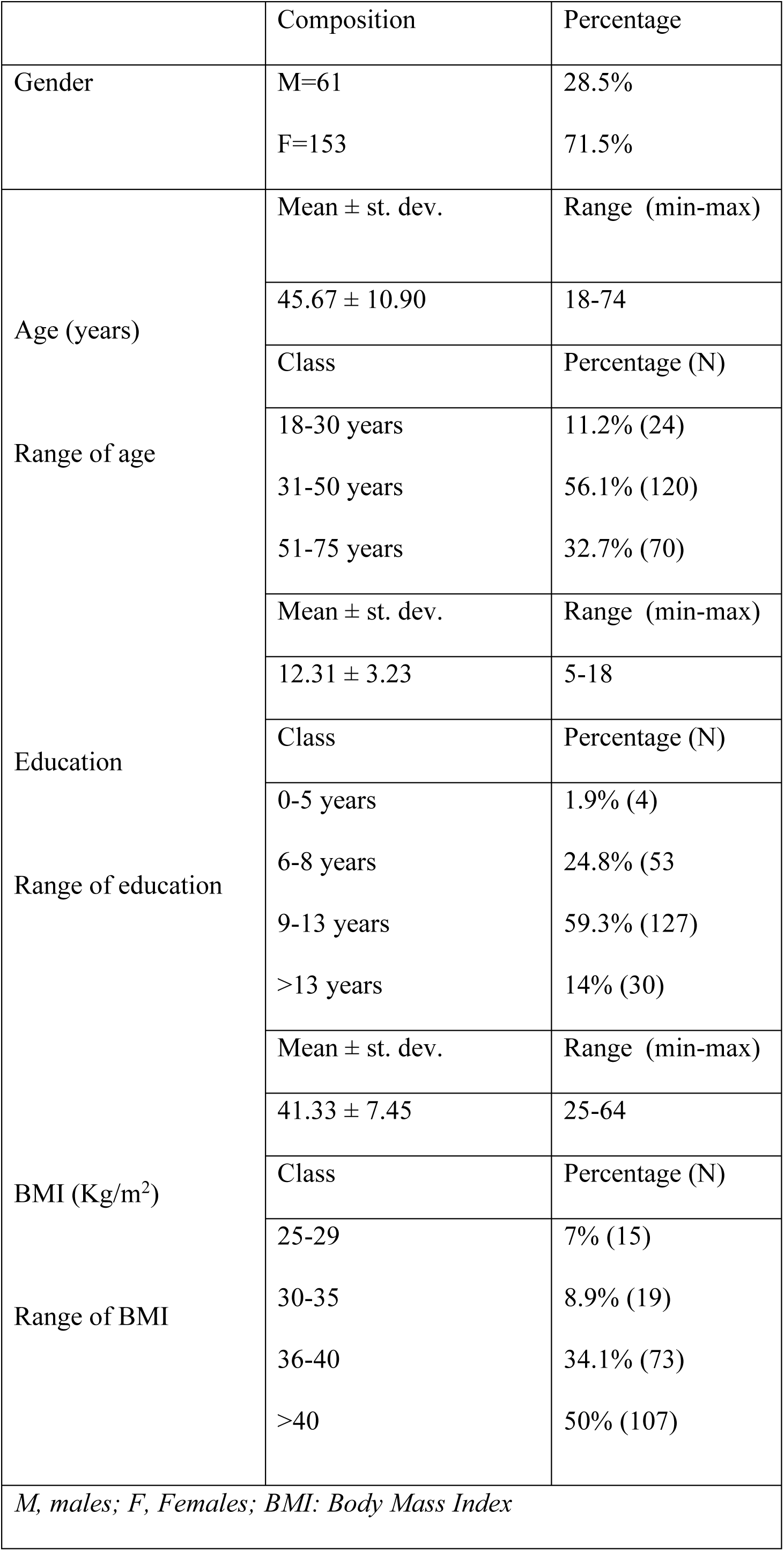
Demographic and descriptive information on the sample.

Concerning the education level, four subjects (1.9%) completed primary school, 53 (24.8%) had a secondary junior school degree, 127 (59.3%) had a high school degree, 30 (14%) had a university degree.

The mean score of each item for the I-WSSQ ranged from 2.03 to 3.78 on the self-devaluation subscale and from 1.94 to 3.48 on the fear of enacted stigma subscale. The skewness of each item for the I-WSSQ ranged from -0.73 to 0.85 on the self-devaluation subscale and from -0.50 to 2.11 on the fear of enacted stigma subscale.

### Reliability

Internal consistency measures for both the subscales and the overall scale were sufficiently good. The internal consistency of the full I-WSSQ was α =.814, and for the subscales—factor 1 and factor 2— internal consistency was α=. 733 and α=. 797, respectively.

### Factor Structure

The mean coefficient correlation among the 12 items was 0.23. However, some correlations resulted low, particularly for items 1 and 2. The Kaiser-Meyer-Oklin value was 0.741, and Bartlett’s test of sphericity reached statistical significance (P<0.00001).

The principal component analysis revealed two components (self-devaluation and fear of enacted stigma) that explained 33.85% and 15.4% of the variance, respectively. Item 1 did not load, as expected, in factor 1 (**Table 2)**. The model fit emerging from confirmatory factor analysis was highly significant (χ^2^ _53_=226.06; p<0.0000001), with factor loading estimates ranging between 0.39 and 1.13.

**Table 2.**
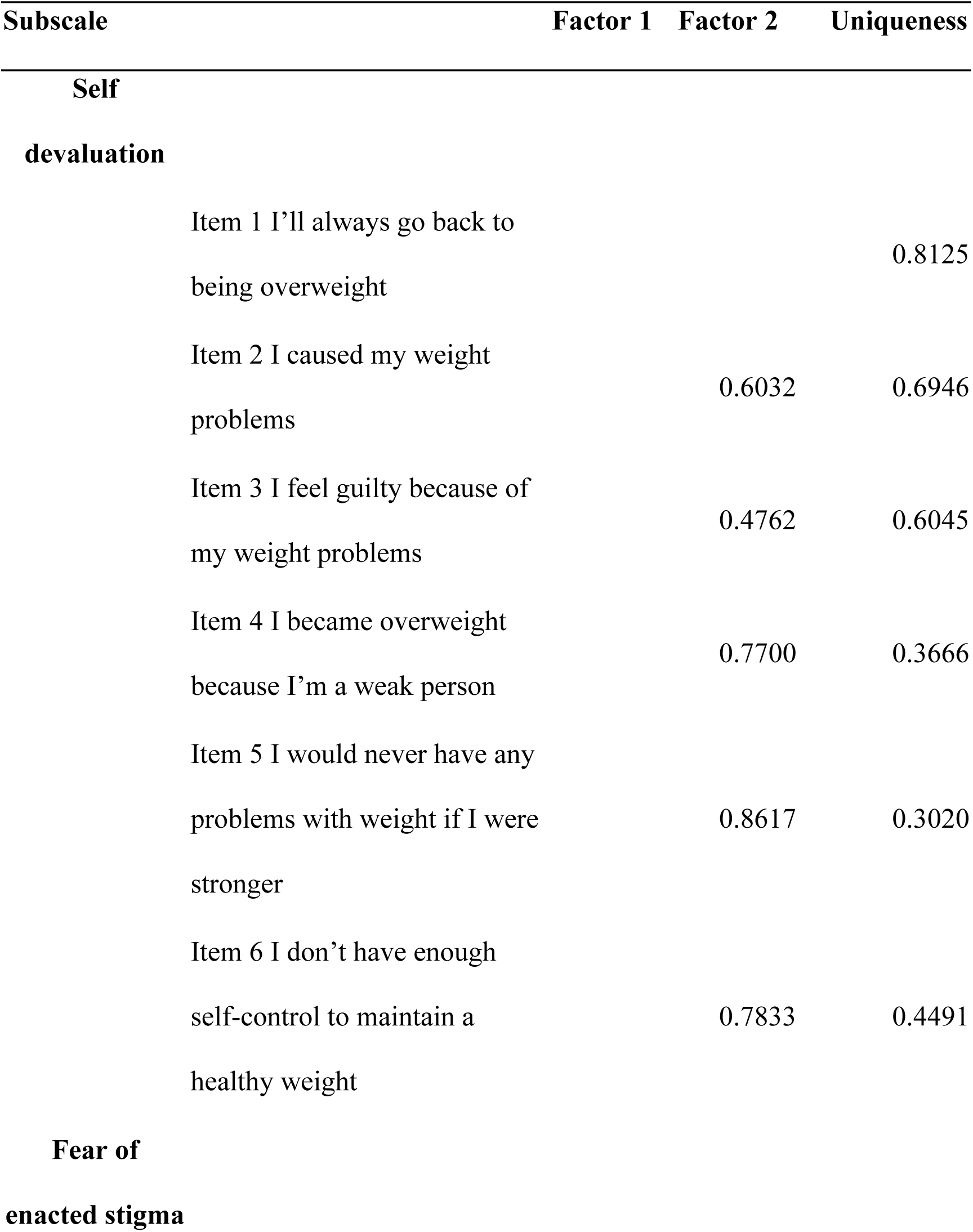
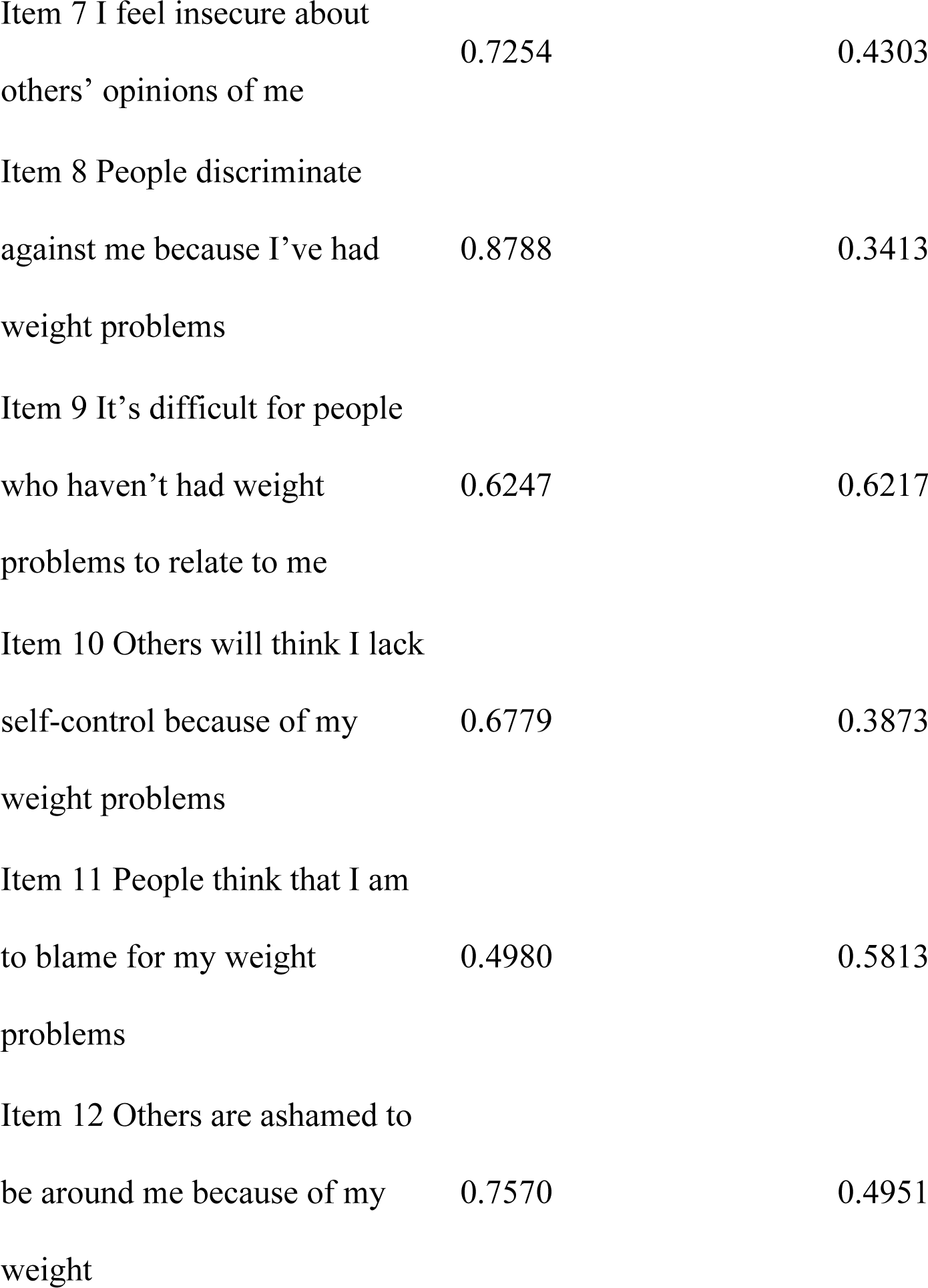
Italian Weight Self-Stigma Questionnaire (I-WSSQ) items: factor loadings from principal components analysis with oblique rotation

| **Subscale** | **Factor 1** | **Factor 2** | **Uniqueness** |
| --- | --- | --- | --- |
| **Self devaluation** |  |  |  |
| Item 1 I'll always go back to being overweight |  |  | 0.8125 |
| Item 2 I caused my weight problems |  | 0.6032 | 0.6946 |
| Item 3 I feel guilty because of my weight problems |  | 0.4762 | 0.6045 |
| Item 4 I became overweight because I'm a weak person |  | 0.7700 | 0.3666 |
| Item 5 I would never have any problems with weight if I were stronger |  | 0.8617 | 0.3020 |
| Item 6 I don't have enough self-control to maintain a healthy weight |  | 0.7833 | 0.4491 |
| **Fear of enacted stigma** |  |  |  |
| Item 7 I feel insecure about others' opinions of me | 0.7254 |  | 0.4303 |
| Item 8 People discriminate against me because I've had weight problems | 0.8788 |  | 0.3413 |
| Item 9 It's difficult for people who haven't had weight problems to relate to me | 0.6247 |  | 0.6217 |
| Item 10 Others will think I lack self-control because of my weight problems | 0.6779 |  | 0.3873 |
| Item 11 People think that I am to blame for my weight problems | 0.4980 |  | 0.5813 |
| Item 12 Others are ashamed to be around me because of my weight | 0.7570 |  | 0.4951 |

The criterion-related validity was examined by performing a correlation analysis between the BMI and scored of BUT A (GSI TOT, global severity index), BUT B (PST, positive symptom total), and I-WSSQ: the Pearson product moment correlation coefficients reported in **Table 3** indicate acceptable criterion-related validity.

**Table 3.** Correlations among WSSQ, BMI, BUT-A (GSI TOT and DISTRESS) and BUT-B (PST and DISTRESS)

| Variable | I-WSSQ | Self-devaluation | Fear of Enacted Stigma |
| --- | --- | --- | --- |
| BMI | 0,0162 | -0,044 | 0,09 |
| BUT-A GSI Tot | 0,501*** | 0.354*** | 0,486*** |
| BUT-B PST | 0,190** | 0,139* | 0,180** |
| I-WSSQ, Italian Weight Self-Stigma Questionnaire, BMI, Body Mass Index, BUT, *Body Uneasiness Test*. GDS TOT, *Global severity index*; PST, *Total positive symptoms*; *p<0,05; **p<0,001; ***p<00001 |  |  |  |

## Discussion

We herein translated and tested the Italian version of the WSSQ for reliability and validity. Compared to the original English version of the WSSQ, with a reported Cronbach α ranging from .812 to .878 (19), the I-WSSQ showed adequate, consistent reliability, comparable to that of the English version, with a Cronbach *α* value of .733 for the fear of enacted stigma subscale (factor 1) and .797 for the self-devaluation subscale (factor 2). Moreover, the inter-item correlation and item-scale correlation analyses revealed that the I-WSSQ had appropriate congruence constructs. The item discrimination analysis indicated that the items in the I-WSSQ had sufficient discriminating power. Furthermore, present results showed that the I-WSSQ had qualitative items to discriminate between the response levels of different participants.

Regarding validity, Lillis et al. reported that the WSSQ fits the 2-factor extraction closely on the self-devaluation and fear of enacted stigma subscales ^35^. In the present study, exploratory factor analysis was used to distinguish between the domain structures of the I-WSSQ and the original English version of the WSSQ, and the domain structures were similar. However, the Italian WSSQ loading of the items was consistent with the original WSSQ, except for item 1 (i.e., “*I’ll always go back to being overweight*”), which did not significantly load in any factor. This result may be explained by the enrolment of some candidates to bariatric surgery, who did not expect to regain weight postoperatively.

Raves and collaborators have shown that internalized stigma and general experience of weight-related stigma predicted poorer diet adherence, even after weight loss in postoperative patients ^38^. While patients do not recognize this correlation, healthcare professionals think it is due to low patients’ compliance. The candidates for bariatric surgery likely expect to lose weight after surgery and never become overweight again. Therefore, this data could be read as a factor of tenacity and resilience. However, more work is needed to understand whether weight stigma plays a role in weight loss outcomes among people undergoing bariatric surgery.

The literature confirms that weight stigma not only exacerbates mental health problems but also hinders physical health maintenance ^25,39^. People with obesity often internalize the weight stigma of others, experiencing psychological distress, including higher body dissatisfaction. This leads to the adoption of behaviours causing weight gain, such as overeating and physical inactivity. It is, therefore, crucial that health professionals assess the weight self-stigma of patients with obesity before any intervention takes place to facilitate the improvement of weight-loss programs ^40-42^.

This is the first study that aimed to translate the WSSQ into Italian. In this study, we performed basic item analysis and internal consistency analysis, and we also conducted a factor analysis and criterion-related correlation analysis. Overall, these results indicate that the I-WSSQ has good reliability and validity and can facilitate verifying the weight self-stigma of people with overweight/obesity whose primary language is Italian.

The I-WSSQ exhibits acceptable reliability and validity and can be applied to people with obesity whose primary language is Italian, allowing to assess self-stigma in this population, with its cultural, psychosocial, and dietary peculiarities. However, the present study has several limitations regarding generalizability. Although the sample size was sufficient to perform factor and correlation analysis, further analysis of the I-WSSQ requires a larger and more heterogeneous sample. Second, the I-WSSQ is a direct translation of the WSSQ, and further research should detect and compare the impact of different cultures on the scale content.

## Conclusions

In conclusion, more attention should be directed to weight-related stigma as a relevant psychosocial factor in obesity-focused prevention and treatment. Therefore, efforts are needed to develop weight stigmatization assessment and measurement tools.

Broadly, this study found that the I-WSSQ has adequate reliability and validity to be used with people with overweight/obesity whose primary language is Italian. Given these acceptable psychometric properties, the I-WSSQ is a valuable tool that enables the rapid assessment of the weight self-stigma within 3 to 5 minutes. Indeed, it could be used to monitor the impact of weight-based stigma and provide helpful information on weight self-stigma to healthcare professionals. Moreover, it may be a reliable tool to explore the issue of self-stigma in different clinical situations, such as weight loss programs involving bariatric surgery, although further *ad hoc* studies are warranted.

## Data Availability

All data produced in the present study are available upon reasonable request to the authors

## Availability of data and material

The datasets generated during and/or analyzed during the current study will be made available upon reasonable request to the corresponding author.

## Author contribution

Conceptualization, LQ, GC, DT; methodology, LQ, GC, MW; formal analysis, GC; data curation, AS FV IB VP, MW; writing—original draft preparation, LQ GC; writing—review and editing, MW DT; supervision, SM, GC, DT ; All authors have read and agreed to the published version of the manuscript.

## Ethics approval

The ethics committee of the University Hospital Campus Biomedico of Rome approved this study and its development. All the procedures performed in the study complied with the ethical standards of the institutional and/or national research committee and the Helsinki Declaration of 1964 and its subsequent amendments or comparable ethical standards. Written informed consent was obtained from all participants prior to starting the study.

